# Severe acute respiratory syndrome coronavirus 2 (SARS-CoV-2) seroprevalence: Navigating the absence of a gold standard

**DOI:** 10.1101/2021.05.11.21256992

**Authors:** Sahar Saeed, Sheila F. O’Brien, Kento Abe, QiLong Yi, Bhavisha Rathod, Jenny Wang, Mahya Fazel-Zarandi, Ashleigh Tuite, David Fisman, Heidi Wood, Karen Colwill, Anne-Claude Gingras, Steven Drews

## Abstract

**Background:** Multiple anti-SARS-CoV-2 immunoassays are available, but no gold standard exists. We assessed four assays using various methodological approaches to estimate SARS-COV-2 seroprevalence during the first COVID-19 wave in Canada.

**Methods:** This serial cross-sectional study was conducted using plasma samples from healthy blood donors between April-September 2020. Qualitative assessment of SARS-CoV-2 IgG antibodies was based on four assays: Abbott Architect SARS-Cov-2 IgG assay (target nucleocapsid) (Abbott-NP) and three in-house IgG ELISA assays (target spike glycoprotein (Spike), spike receptor binding domain (RBD), and nucleocapsid (NP)). Seroprevalence was estimated using multiple composite reference standards (CRS) and by a series of Bayesian Latent Class Models (BLCM) (using uninformative, weakly, and informative priors).

**Results:** 8999 blood samples were tested. The Abbott-NP assay consistently estimated seroprevalence to be lower than the ELISA-based assays. Discordance between assays was common, 13 unique diagnostic phenotypes were observed. Only 32 samples (0.4%) were positive by all four assays. BLCM using uninformative priors predicted seroprevalence increased from 0.7% (95% credible interval (CrI); 0.4, 1.0%) in April/May to 0.8% (95% CrI 0.5, 1.2%) in June/July to 1.1% (95% CrI 0.7, 1.6) in August/September. Results from CRS were very similar to the BLCM. Assay characteristics varied considerably over time. Overall spike had the highest sensitivity (89.1% (95% CrI 79.2, 96.9%), while the sensitivity of the Abbott-NP assay waned from 65.3% (95% CrI 43.6, 85.0%) in April/May to 45.9% (95% CrI 27.8, 65.6) by August/September.

**Discussion:** We found low SARS-CoV-2 seroprevalence rates at the end of the first wave and estimates derived from single assays may be biased.

**Summary:** Multiple anti-SARS-CoV-2 immunoassays are available, but no gold standard exists. We used four unique assays to estimate very low SARS-COV-2 seroprevalence during the first COVID-19 wave in Canada. Caution should be exercised when interpretating seroprevalence estimates from single assays.

## INTRODUCTION

Worldwide, more than 155 million people have been diagnosed with coronavirus disease 2019 (COVID-19), as of May 7, 2021 [1]. Yet, this is likely an underestimation of the true burden of severe acute respiratory syndrome coronavirus 2 (SARS-CoV-2) given testing is primarily used to confirm suspected infection as opposed to broad surveillance. For example, in Canada, testing was only accessible early in the pandemic to people who had symptoms, were known contacts of a case or had a relevant travel history [2]. This meant community transmission by asymptomatic or mildly symptomatic individuals was likely underestimated. Determining the proportion of individuals with evidence of an immune response to SARS-CoV-2 can provide a more comprehensive assessment of prevalence to assist public health officials in making policy decisions. This prompted an urgent need for seroprevalence studies and accurate anti-SARS-CoV-2 immunoassays to detect the true burden of disease.

While multiple commercial and in-house immunoassays to detect anti-SARS-CoV-2 antibodies are available, to date no gold standard exists [3]. Furthermore, laboratorians have described multiple examples of discordance between assays [4]. Some of this variability is in part due to the assays which vary significantly by the isotype (i.e. IgA, IgM, IgG), viral antigens (i.e. spike or nucleocapsid protein and whether full-length or partial), and test performance (i.e. sensitivity/specificity). It is also known that anti-SARS-COV-2 antibodies wane over time which can further affect the sensitivity and specificity of the assays [5–7]. Additionally, biological differences between individuals can lead to different antibody profiles. Given these overlapping challenges of estimating seroprevalence, relying on a single assay (regardless which assay this may be) may bias results.

In the absence of a gold standard, using results from multiple assays may improve accuracy. However, which methods are appropriate for estimating SARS-CoV-2 seroprevalence has not been defined. One method is to use a composite reference standard (CRS); a traditional approach used in clinical settings based on prespecified rules from orthogonal testing [8]. More recently, Bayesian Latent Class Analysis (BLCA) has become more mainstream in diagnostic studies [9]. In contrast to CRS which classifies individuals as either positive or negative, BLCA uses a likelihood-based approach from multiple imperfect assays to estimate test characteristics and prevalence. Given the uncertainty of the assay performance, we evaluated multiple methodological approaches to estimate SARS-COV-2 seroprevalence during the first COVID-19 wave in Canada using four unique assays.

## METHODS

### Study Design and Population Sampling

We conducted a serial cross-sectional study among blood donors in Canada between April and September 2020 (Prior to COVID-19 vaccine availability). Canadian Blood Services collects approximately 850,000 blood donations per year from a combination of fixed and mobile sites in all larger cities and most urban areas from all provinces in Canada except Quebec [10]. Blood donors (>17 years old) must meet numerous selection criteria to ensure that they are in good health and at low risk of infectious disease. Beginning in March 2020, donors were deferred for two weeks if they were diagnosed with SARS-CoV-2 infection or if they were in contact with a known case. Each month 1500 deidentified samples were randomly selected by collection site. Data on the collection site, birth year, sex and Forward Sortation Area (FSA) of the residential postal code for each donor were extracted. The Research Ethics Board of the Canadian Blood Services and Lunenfeld-Tanenbaum Research Institute (LTRI) (REB study #20-0194-E) approved this study and exempted study-specific consent.

### SARS-CoV-2 antibody testing

Retention EDTA plasma samples were aliquoted and frozen at −20°C at the CBS laboratory in Ottawa. Each sample was tested for SARS-CoV-2 IgG antibodies using four assays. The Abbott Architect SARS-Cov-2 IgG assay which targets the nucleocapsid antigen (Abbott-NP), (Abbott, Chicago IL) and three in-house IgG ELISA chemiluminescent assays recognizing distinct recombinant viral antigens: full length spike glycoprotein (Spike), spike glycoprotein receptor binding domain (RBD), and nucleocapsid (NP), were tested at the CBS laboratory in Ottawa and the Gingras laboratory [11,12] at the LTRI in Toronto, respectively. Table 1 summarizes each antibody assay by: platform, antigen targets and how reactivity was determined.

**Table 1.** Assay Characteristics.

| Assay | Assay platform | Capture Antigen (IgG) | Manufacture | Cut-offs (positive) | Cut-off reference* |
| --- | --- | --- | --- | --- | --- |
| Abbott-NP | Chemiluminescent microparticle immunoassay | Nucleocapsid | Abbott | $\geq 1.40$ | Manufacture |
| Spike | Chemiluminescent ELISA | spike | Gingras Lab | $\geq 0.190$ | 3 SD + negative mean |
| RBD | Chemiluminescent ELISA | RBD | Gingras Lab | $\geq 0.186$ | 3 SD + negative mean |
| NP | Chemiluminescent ELISA | Nucleocapsid | Gingras Lab | $\geq 0.396$ | 3 SD + negative mean |

\*3 Standard deviations (SD) + negative mean is a standard approach to choosing cut-off thresholds for ELISA based assays. Briefly, the relative ratio values of the negative controls from 20-22 different tests were used. Ratios are transformed on the log10-scale and then mean 3 SD determines the cutoff. Values are then exponentiated to identify cut off listed in the table.

### Analysis

We evaluated the correlation between the individual assays by kappa statistics. In the absence of a gold standard, we examined multiple approaches to estimate seroprevalence. First, seroprevalence was estimated by individual assays based on pre-defined thresholds (Table 1). Then we used a series of composite reference standards to identify a “true” positive if a sample was reactive by a combination of two or more assays. Finally, we evaluated seroprevalence a Bayesian latent class analysis.

#### Bayesian Latent Class Analysis (BLCA)

In this study the “latent” unobservable target was evidence of SARS-CoV-2 infection (based on IgG positivity). Instead of relying on one imperfect assay, this iterative model leverages the data from multiple imperfect assays to estimate the “true” prevalence and test characteristics [13,14]. Given any one of four assays could assign an individual to be positive or negative, there was a maximum of 16 (2^4^), possible diagnostic phenotypes. For this model, we assumed each assay was independent of the others, conditional on the individual’s unknown antibody status. This means that the probability of obtaining a given diagnostic phenotype depended on the probability that an individual had been truly infected with SARS-CoV-2 and on the outcome of each assay given the underlying exposure status. Briefly, we estimated parameters in a Bayesian framework using a Gibbs sampler to produce Markov chain Monte Carlo (MCMC) simulations. We ran 50,000 iterations, with the first 5000 steps discarded as burn-in. Given the uncertainty of assay performance in a “healthy” population, we used uninformative priors (uniform distributions). As a sensitivity analysis, we did evaluate “informative priors” based on +/-5% assumed sensitivity and +/-1% assumed specificity [12]. We also evaluate the model using “weakly informative priors” (sensitivity (range of 60%-100%) and specificity (range of 90%-100%), for each assay. We verified convergence of all MCMC chains. We reported posterior means and 95% credible intervals (CrI) for all estimated parameters overall and by two-month intervals using SAS (version 9.1, SAS Institute, Cary, NC).

## RESULTS

Between April and September 2020, a total of 8999 healthy blood samples were assessed for SARS-CoV-2 antibodies by four distinct assays. Most donors (96%) were between 20-69 years old, there were slightly more male donors (52%) compared to female donors (48%) and there was representation from all provinces across Canada except Quebec. Donor characteristics remained consistent over the study period (**Supplemental Table 1**).

### Individual Assays

We evaluated seroprevalence rates over time by the individual assays (**Figure 1**). The Abbott-NP assay consistently remained lower than the ELISA-based assays. Seroprevalence based on the spike assay was 3.1% in May, dropped to 1.2% in July and then plateaued around 3%. Rates were lower and more stable by RBD that started at 0.8% and increased to 1.6% by September. In contrast the NP assay increased significantly from May (1.2%) until June (3.7%). The signal to cut off ratios remained relatively stable over time for all assays (**Supplemental Figure 1**). Overall, the kappa score was low (0.28 p<0.0001) and the percent agreement was highest between Abbott-NP and RBD (99% agreement; kappa 0.43).

**Figure 1.**
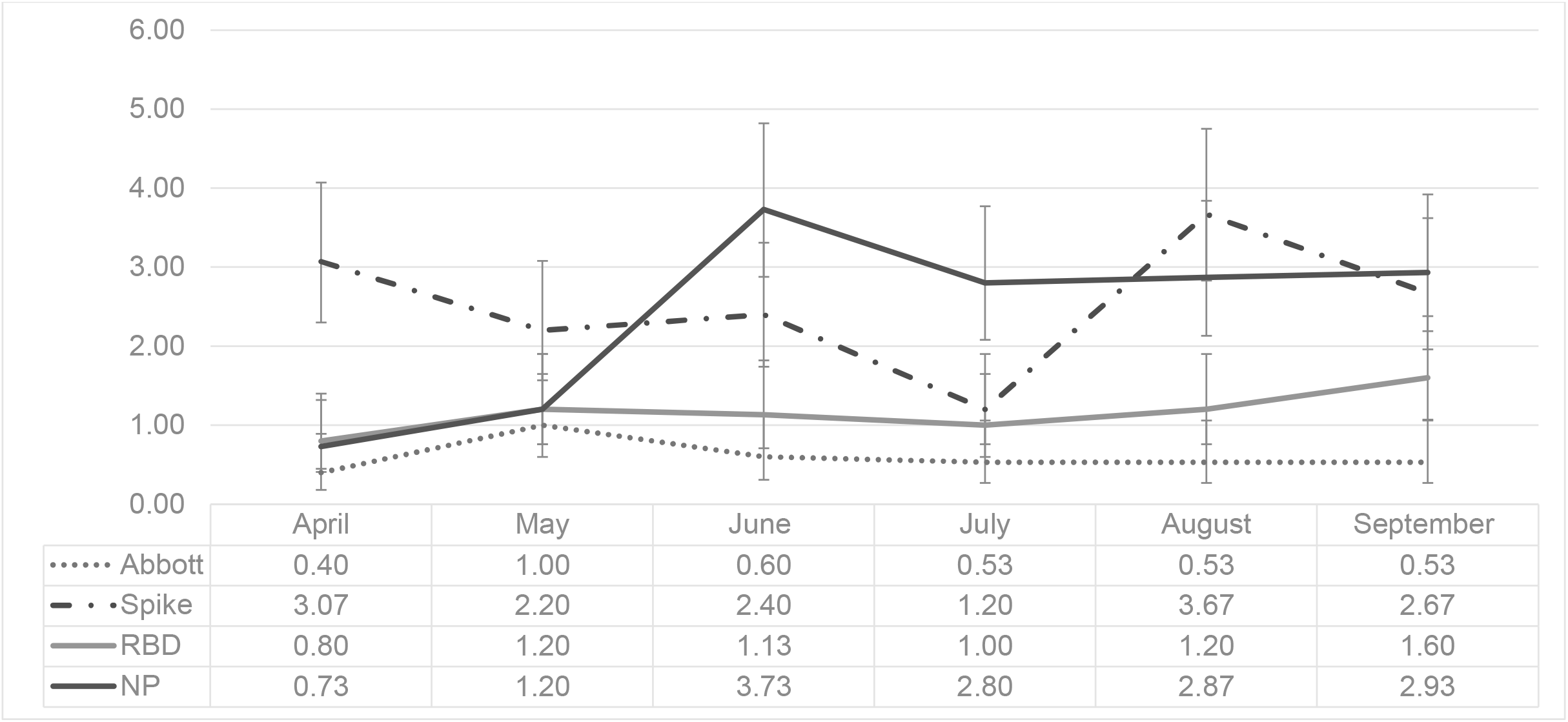
Seroprevalence of the single assay by month over the first COVID-19 wave in Canada. Each line represents seroprevalence rates (summarized in table below) monthly between April and September 2020 (during the first COVID-19 Wave) based thresholds for each assay. Abbott Architect SARS-Cov-2 IgG assay (Abbott-NP) and three in-house IgG ELISA assays recognizing distinct recombinant viral antigens: full length spike glycoprotein (Spike), spike glycoprotein receptor binding domain (RBD), and nucleocapsid (NP).

### Orthogonal Approach

Given screening occurred in a low prevalence setting, to minimize false positive results, we assumed a true positive was more likely when two or more assays were positive. *A priori*, choosing two pre-specified assays resulted in a range of seroprevalence estimates that ranged from 0.2% to 0.5% in April to 0.4% to 1.4% in September (**Figure 2**). Any two assays (from four) resulted in a seroprevalence that increased significantly over time from 0.5% (95% CI 0.3%, 1.1%) in April to 1.3% (95% CI 0.8, 2.0) in September (p=0.02) (**Figure 2**).

**Figure 2.**
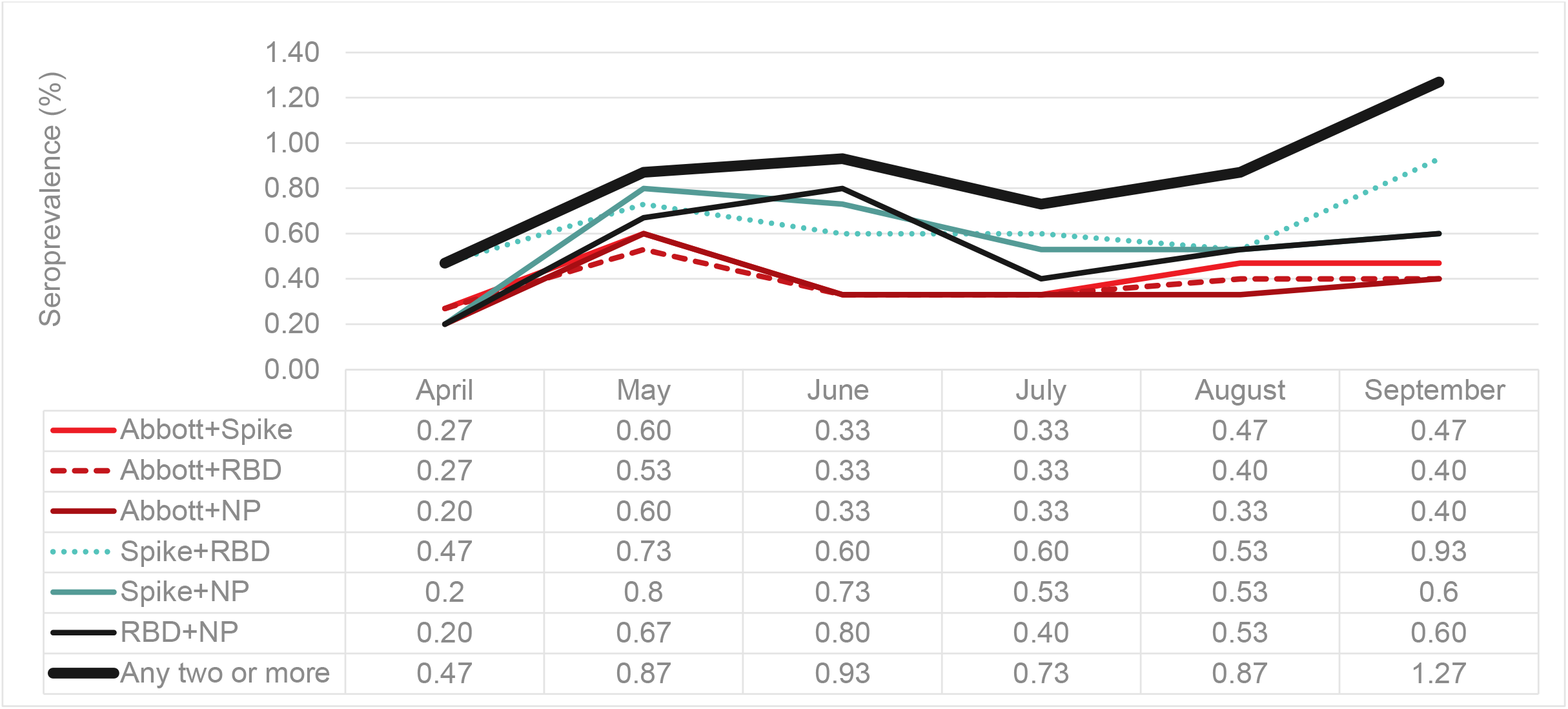
Orthogonal analysis of multiple anti-SARS-CoV-2 immunoassays. Each line represents seroprevalence rates (summarized in table below) monthly between April and September 2020 (during the first COVID-19 Wave) based on predefined definitions. Since we are not comparing CRS, we did not include 95% CI for each data point (all overlapping). CRS based on a combination of reactive samples using: Abbott Architect SARS-Cov-2 IgG assay (Abbott-NP) and three in-house IgG ELISA assays recognizing distinct recombinant viral antigens: full length spike glycoprotein (Spike), spike glycoprotein receptor binding domain (RBD), and nucleocapsid (NP). Positivity based on “any two or more” was determined by a reactive sample from two or more assays)

### Bayesian Latent Class Model

From 16 possible diagnostic phenotypes, 13 were observed among the 8999 sampled (Eight phenotypes had at least 10 observations). The most frequent profile was “all negative” (95.0% (95% CI 94.6, 95.5) followed by only positive by the individual ELISA-based assays (Spike only (1.8%, 95%CI 1.5, 2.0) and NP only (1.7, 95%CI 1.5, 2.1). Only 32 samples were positive for all four assays (0.4, 95%CI 0.3, 0.5) (**Table 2**). Overall seroprevalence was estimated to be 0.8% (95% CrI 0.6, 1.0%); 0.8% (95% CrI 0.6, 1.0%); 0.8% (95% CrI 0.7, 0.9%) using informative, weakly informative and non-informative priors, respectively. **Figure 3** illustrates temporal trends in seroprevalence by BLCA comparing the different priors. The model with the non-informative prior consistently was higher than the other two models, but the difference was not statistically significant. Given the uncertainty of test characteristics, we compared the observed vs predicted values of the three BLCM and found the uninformative model identified the all negative phenotype most accurately.

**Table 2.** Diagnostic Phenotypes of anti-SARS-CoV-2 immunoassay results from 8999 samples tested.

|  | Abbott<br>N=54 | In house ELISA |  |  | Observed |  |
| --- | --- | --- | --- | --- | --- | --- |
|  |  | Spike<br>N=228 | RBD<br>N=104 | NP<br>N=214 | # | % (95% CI) |
| **NONE** | - | - | - | - | 8557 | 95.0 (94.6, 95.5) |
| **ALL** | + | + | + | + | 32 | 0.4 (0.3, 0.5) |
| **All – Abbott** | - | + | + | + | 9 | 0.1 (0.1, 0.2) |
| **NP Only** | - | - | + | - | 156 | 1.7 (1.5, 2.1) |
| **RBD Only** | - | - | - | + | 39 | 0.4 (0.3, 0.6) |
| **Spike Only** | - | + | - | - | 158 | 1.8 (1.5, 2.0) |
| **Spike+RBD** | - | + | + | - | 15 | 0.2 (0.1, 0.3) |
| **RBD+NP** | - | - | + | + | 7 | 0.1 (0.0, 0.3) |
| **Abbott Only** | + | - | - | - | 17 | 0.2 (0.1, 0.3) |
| **Spike+Abbott** | + | + | - | - | 2 | 0.0 (0.0, 0.1) |
| **Spike+NP** | - | + | - | + | 9 | 0.1 (0.1, 0.2) |
| **All –NP** | + | + | + | - | 2 | 0.0 (0.0, 0.1) |
| **All-RBD** | + | + | - | + | 1 | 0.0 (0.0, 0.1) |

Abbott- Architect SARS-Cov-2 IgG assay which targets the nucleocapsid antigen and three in-house IgG ELISA chemiluminescent assays recognizing distinct recombinant viral antigens: full length spike glycoprotein (Spike), spike glycoprotein receptor binding domain (RBD), and nucleocapsid (NP).

**Figure 3.**
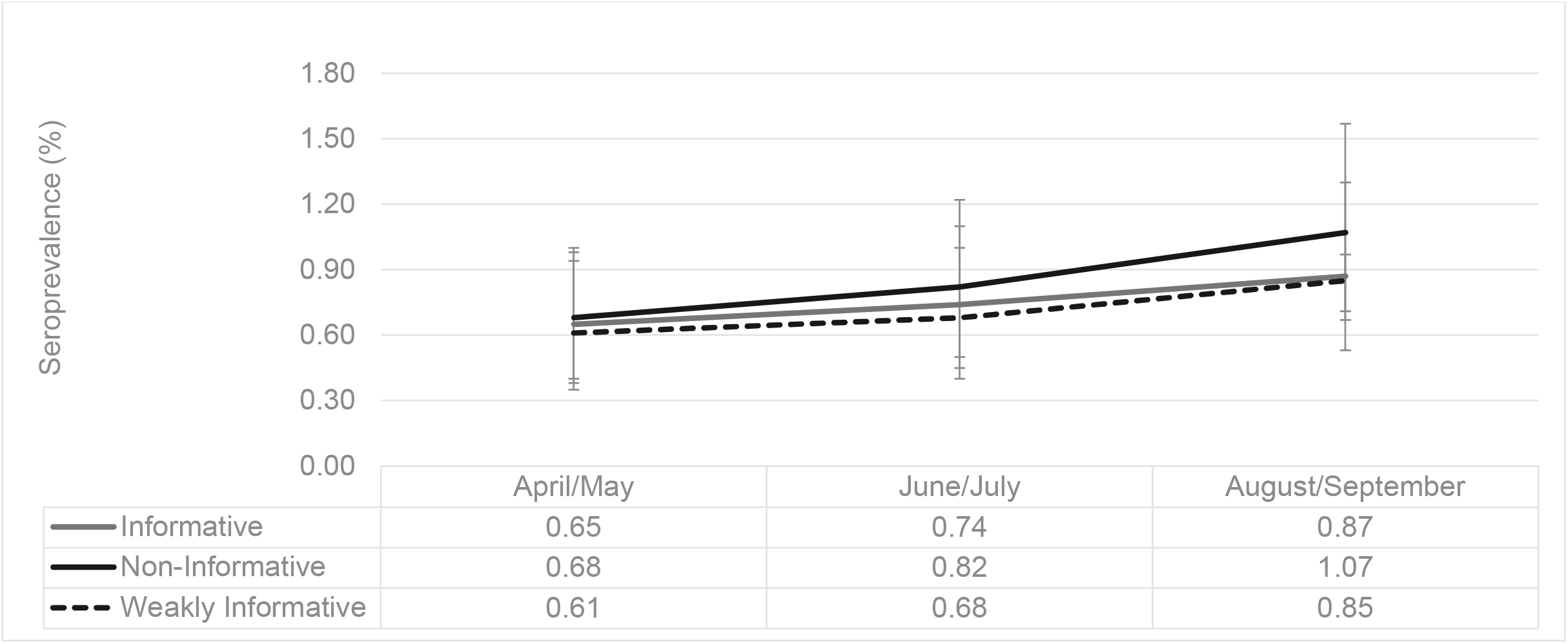
Seroprevalence estimates by different BLCM (Informative, Weakly informative and Non-informative priors) Each line represents seroprevalence rates (summarized in table below) derived from posterior means of three BLCMs (comparing informative, weakly informative and non-informative priors) bi-monthly between April and September 2020 (during the first COVID-19 Wave). Error bars represent 95% Credible Intervals.

The test characteristics (sensitivity and specificity) varied significantly by the different assays **(Table 3)**. Overall, the ELISA based assays had higher sensitivity than the Abbott-NP. RBD had the highest specificity (99.5% (95% CrI 99.4, 99.7%)) and NP had the lowest specificity (98.2% (95% CrI 97.9, 98.4%)). Abbott-NP had a sensitivity of 51.6% (95% CrI 39.4, 64.2%) and a specificity of 99.8% (95% CrI 99.7, 99.9%). Negative predictive values of all assays were very high (ranging from 99.6% to 99.9%). The Abbott-NP had the highest positive predictive value at 67.9% (95% CrI 64.8, 70.4%) while the ELISA based assays that ranged from 24.2 to 60%. Test characteristics varied considerably over time as the first wave of the pandemic progressed (**Table 3**). Sensitivity Abbott-NP assay waned the most from 65.3% (95%CrI 43.6, 85.0) in April/May to 45.9% (95% CrI 27.8, 65.6). Similar trends were observed using informative and non-informative priors (**Supplemental Table 2 and 3**).

**Table 3.** Assay Characteristics overall and over time; based on the BLCM Posterior Means (95% Creditable Interval) (non-informative priors)

|  | **Overall**<br>**Prevalence: 0.80% (95%CrI 0.73, 0.87)** |  |  |  | **April/May**<br>**0.68% (0.57, 0.78)** |  | **June/July**<br>**0.82% (0.69, 0.93)** |  | **August/September**<br>**1.08% (0.91, 1.21)** |  |
| --- | --- | --- | --- | --- | --- | --- | --- | --- | --- | --- |
|  | Sensitivity | PPV | Specificity | NPV | Sensitivity | Specificity | Sensitivity | Specificity | Sensitivity | Specificity |
| Spike | 89.1%<br>(79.2, 96.9) | 28.3%<br>(25.9, 31.1) | 98.2%<br>(97.9, 98.4) | 99.9%<br>(99.9, 100.0) | 92.2%<br>(77.2, 99.4) | 97.9%<br>(97.4, 98.4) | 82.5%<br>(63.6, 96.3) | 98.8%<br>(98.4, 99.2) | 81.9%<br>(63.4, 96.2) | 97.6%<br>(97.0, 98.2) |
| RBD | 86.3%<br>(75.0, 95.3) | 60.0%<br>(54.8, 64.4) | 99.5%<br>(99.4, 99.7) | 99.9%<br>(99.8, 100.0) | 84.2%<br>(65.4, 96.7) | 99.5%<br>(99.2, 99.7) | 82.7%<br>(62.6, 97.3) | 99.6%<br>(99.3, 99.8) | 78.5%<br>(59.0, 93.9) | 99.4%<br>(99.0, 99.7) |
| NP | 71.8%<br>(60.1, 82.5) | 24.2%<br>(22.0, 26.6) | 98.2%<br>(97.9, 98.4) | 99.8%<br>(99.7, 99.8) | 72.6%<br>(51.3, 90.4) | 99.5%<br>(99.2, 99.7) | 81.0%<br>(62.3, 94.7) | 97.3%<br>(96.7, 97.9) | 58.1%<br>(39.3, 76.2) | 97.6%<br>(97.0, 98.2) |
| Abbott-NP | 51.6%<br>(39.4, 64.2) | 67.9%<br>(64.8, 70.4) | 99.8%<br>(99.7, 99.9) | 99.6%<br>(99.5, 99.7) | 65.3%<br>(43.6, 85.0) | 99.7%<br>(99.5, 99.9) | 43.8%<br>(24.8, 64.6) | 99.7%<br>(99.5, 99.9) | 45.9%<br>(27.8, 65.6) | 99.9%<br>(99.7, 100.0) |

Abbott- Architect SARS-Cov-2 IgG assay which targets the nucleocapsid antigen and three in-house IgG ELISA chemiluminescent assays recognizing distinct recombinant viral antigens: full length spike glycoprotein (Spike), spike glycoprotein receptor binding domain (RBD), and nucleocapsid (NP)

**Figure 4** compares seroprevalence rates based on the three methods (>2 reactive assays, BLCA-non-informative priors) and compares to the Abbott-NP (a common commercial assay). The latent class model and orthogonal approach yielded almost identical results and there was no evidence of waning seroprevalence rates over time.

**Figure 4.**
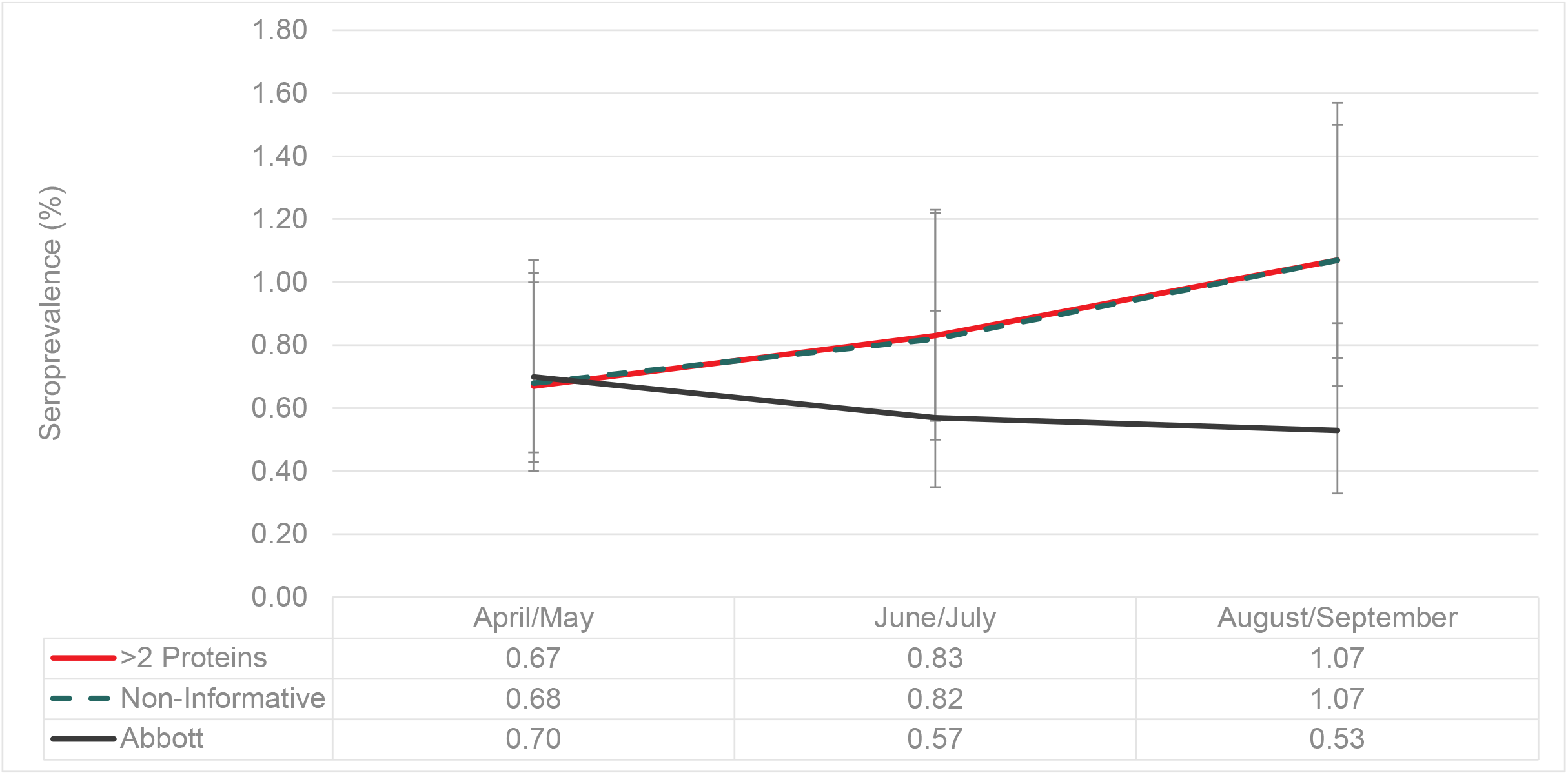
Summary comparison of seroprevalence rates by analytical methods. Each line represents seroprevalence rates (summarized in table below) derived from four analytical methods bi-monthly between April and September 2020 (during the first COVID-19 Wave). >2 proteins (positivity was determined by a reactive sample from two or more assays), BLCA-Bayesian latent class analysis with non-informative priors, results are posterior means and error bars are 95% CrI and Abbott-NP is a single commercial assay.

## DISCUSSION

In the absence of a gold standard, we evaluated multiple assays and methodological approaches to estimate SARS-CoV-2 seroprevalence in healthy Canadian blood donors. Results from orthogonal approaches varied by how many assays were chosen a priori. Using either a latent class analysis or a composite reference standard of at least two positive assays (out of four) generated similar results. Regardless of the analytical method, SARS-CoV-2 seroprevalence was very low (~1%) at the end of the first COVID-19 wave among a healthy population of blood donors. This may be due in part to early travel and social contact restrictions that were widely implemented in multiple Canadian jurisdictions.

Gaps in laboratory testing during the first wave, the significant proportion of asymptomatic or pauci-symptomatic infections, as well as a continuing pandemic have prompted public health authorities in Canada to continue to invest in serological surveys to evaluate the true burden of SARS-CoV-2. Yet unique biological and epidemiological challenges exist when estimating seroprevalence, particularly in low prevalence settings. We recently conducted a scoping review and identified 33 seroprevalence studies among blood donors worldwide. From the 33 studies, 27 unique assay combinations were identified, more than half of studies used a single assay to determine prevalence and less than a third accounted for imperfect test performance [15]. Results from this study suggest relying on a single assay to determine prevalence in a low prevalence setting may significantly bias results.

The variability in the number of diagnostic phenotypes may be associated with the interindividual variability of the immune response. SARS-CoV-2 infects cells using a spike glycoprotein to bind to human angiotensin-converting enzyme 2 (ACE2) [16–18]. The receptor binding domain attached to spike mediates both viral binding and fusion events and all proteins are targets for neutralizing monoclonal antibodies [19,20]. Biologically, it is not clear why a person may differentially express antibodies against SARS-CoV-2, but among our sample, we found 13 distinct diagnostic phenotypes. The discordance between assays may also be a product of imperfect test characteristics. In this study, we used one commercial assay for which the manufacture originally reported a sensitivity of 95.9% and specificity of 99.6%. Later real-world reports suggested the sensitivity was as low as 92.7% [21–26]. Results from this study suggest significantly lower sensitivity. While it is customary to assume that assay performance remains static, amid this dynamic pandemic, waning antibody signals may compromise correct classification of prior SARS-CoV-2 exposure. We have previously shown in a longitudinal study that the NP signal in the ELISA-based assay wanes faster than spike or RBD [12]. Consistent with previous reports, we found the nucleocapsid signal from the Abbott assay also wanes faster than spike or RBD [27,28]. This suggests that NP-based assays may be identifying more recent exposures.

It should be noted that waning antibody signals do not necessarily mean waning cellular mediated immunity. Indeed, recent studies suggest in the absence of detectable antibody signals there is evidence of neutralization associated with longer lasting immunity [29,30]. Therefore, without adjusting for waning antibody signals we may be underestimating SARS-CoV-2 seroprevalence. At this point in time, it remains unknown what the true measures or correlates of immunity are in the Canadian population. The data presented here does not address whether some blood donors may have mounted a cellular immune response with an antibody response that waned by the time of serologic testing. We also note that the presence of antibodies does not imply that those antibodies are neutralizing; although we have assessed for spike and RBD antibodies, we have not attempted to understand the neutralizing capacity of these donor specimens against wild type strains of SARS-CoV-2 or emerging variants in Canada. In the next steps of our analysis we will be undertaking studies to understand the neutralizing capacity of these donor specimens to SARS-CoV-2.

Our study has several strengths. First this study is nested within a large national seroprevalence survey which to date has tested >179,000 samples using the Abbott-NP assay since the beginning of the pandemic in Canada. While we tested only a fraction of the samples, seroprevalence rates (based on Abbott-NP) were very similar, illustrating the generalizability of our results nationally [31]. Given the uncertainty around the assay characteristics specifically among an asymptomatic population, we used multiple methodological approaches to estimate seroprevalence and report all findings. One of the strengths of the BLCA is the ability to estimate assay performance. Given limited resources, it may not be feasible to evaluate seroprevalence using four unique assays. However, smaller nested studies with more comprehensive antibody data within larger surveys can be used to correct for measurement errors. For example, we used the sensitivity and specificity of the BCLA from this study to adjust for national seroprevalence estimates recently published between May-July 2020 [31]. We reported seroprevalence was 0.74% (95%CI 0.68, 0.80), but after reanalyzing the data with updated sensitivity/specificity, based on the BLCM with non-informative priors, we found that the corrected seroprevalence was 47% higher at 1.03% (95% CI 0.91, 1.14%). As the pandemic continues, the proportion of recent and older infections will continue to vary over time and having an ability to correct these time varying assay characteristics will become even more important.

Our study also has weaknesses. This study was conducted among blood donors, based on selection criteria to be allowed to donate blood donors may be healthy than the general population [32]. However, a recent study compared seroprevalence estimates from European blood donors to household surveys targeting the general population and found seroprevalence rates to be very similar [33]. Both CRS and LCA assume each of the assays is conditionally independent. It is possible this assumption may not hold, potentially biasing results. Assay performance is based on predefined thresholds. For the Abbott assay we used the manufacturer’s ≥1.4 cut off, but recent reports do suggest reducing the threshold to >0.8 to increase sensitivity and to account for waning antibody signals. However, the sensitivity and specificity was not available by the manufacture for us to evaluate this alternative threshold. All four assays only probed for IgG meaning that we did not measure IgM and IgA, which may provide some neutralizing capacity in some individuals as anti-SARS-CoV-2 IgG titers begin to rise. In other donors, different profiles of anti-SARS-COV-2 IgM, IgA and IgG may also provide different profiles of humoral protection. Finally, we did not assess for the SARS-CoV-2 neutralizing capacity of donor specimens nor the avidity of the IgG antibody responses in those donors.

In conclusion, regardless of the analytical method we found at the end of the first COVID-19 wave, SARS-CoV-2 seroprevalence among a healthy population of Canadian blood donors was low (~1%) and the sensitivity of all the assays waned. This is supported by low rates of SARS-CoV-2 case detections reported nationally within this time-period [34]. These findings suggest significant limitations to using a single assay to estimate SARS-CoV-2 seroprevalence in a low prevalence setting, such as healthy Canadian blood donors during the first wave of the COVID-19 pandemic. We recommend seroprevalence studies to use multiple assays on either their entire sample or at least a subset to estimate seroprevalence more accurately in the future.

## Data Availability

Data is propriety

## Acknowledgements

We would like to thank Craig Jenkins, Valerie Conrod and Canadian Blood Services operations staff for their assistance with this project.

