## Supplemental Tables and Figures for "Severe acute respiratory syndrome coronavirus 2 (SARS-CoV-2) seroprevalence: Navigating the absence of a gold standard"

### Supplemental material

**Supplemental Figure 1.** Signal to cut off ratio (S/Co) by calendar month April (4) until September (9). Red lines represent thresholds. Abbott-NP (1.4) (n=54 positive) based on the manufacture's recommendations. Spike (0.190) (n=228 positive); RBD (0.186) (n=104 positive); and NP (0.396) (n=214 positive). Abbott- Architect SARS-Cov-2 IgG assay which targets the nucleocapsid antigen and three in-house IgG ELISA chemiluminescent assays recognizing distinct recombinant viral antigens: full length spike glycoprotein (Spike), spike glycoprotein receptor binding domain (RBD), and nucleocapsid (NP).

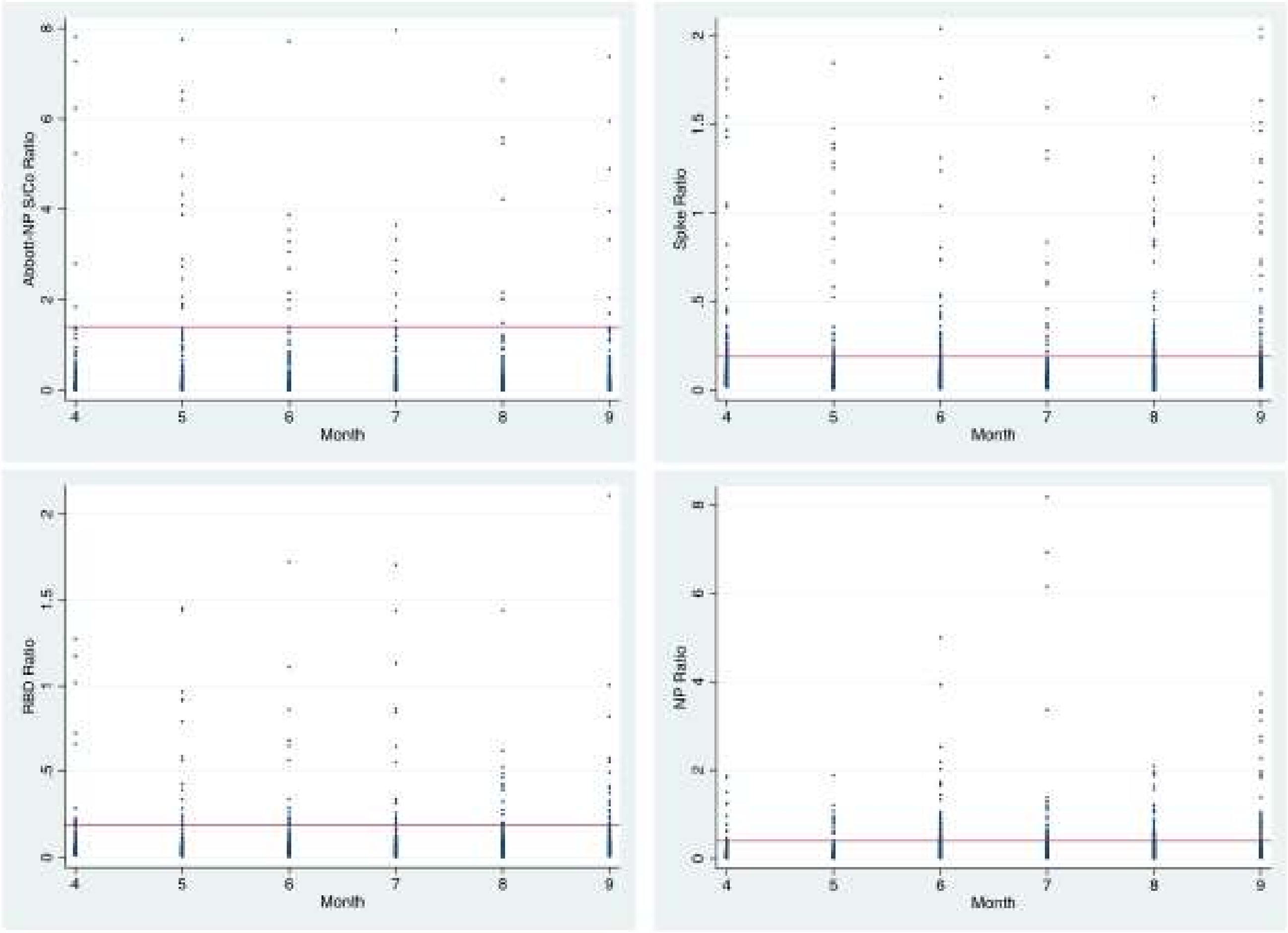

**Supplemental Table 1. Baseline Characteristics of blood donor populations over time**

| | | **Overall** | | April | May | June | July | Aug | Sept |
|---|---|---|---|---|---|---|---|---|---|
| Province | | | | | | | | | |
| | British Columbia | **1358** | **15%** | 228 (15%) | 228 (15%) | 227 (15%) | 229 (15%) | 228 (15%) | 218 (14%) |
| | Alberta | **1862** | **21%** | 309 (21%) | 309 (21%) | 309 (21%) | 308 (21%) | 308 (21%) | 319 (21%) |
| | Saskatchewan | **439** | **5%** | 77 (5%) | 54 (4%) | 77 (5%) | 77 (5%) | 77 (5%) | 77 (5%) |
| | Manitoba | **496** | **6%** | 79 (5%) | 102 (7%) | 80 (5%) | 78 (5%) | 78 (5%) | 79 (5%) |
| | Ontario | **3889** | **43%** | 648 (43%) | 647 (43%) | 648 (43%) | 649 (43%) | 649 (43%) | 648 (43%) |
| | Atlantic | **955** | **11%** | 159 (11%) | 159 (11%) | 159 (11%) | 159 (11%) | 160 (11%) | 159 (11%) |
| Age group | | | | | | | | | |
| | 17-19 | **185** | **2%** | 23 (2%) | 19 (1%) | 36 (2%) | 33 (2%) | 44 (3%) | 30 (2%) |
| | 20-29 | **1525** | **17%** | 253 (17%) | 248 (17%) | 240 (16%) | 232 (15%) | 286 (19%) | 266 (18%) |
| | 30-39 | **1655** | **18%** | 286 (19%) | 270 (18%) | 284 (19%) | 255 (17%) | 244 (16%) | 316 (21%) |
| | 40-49 | **1453** | **16%** | 249 (17%) | 247 (16%) | 207 (14%) | 233 (16%) | 271 (18%) | 246 (16%) |
| | 50-59 | **1895** | **21%** | 336 (22%) | 319 (21%) | 338 (23%) | 333 (22%) | 292 (19%) | 277 (18%) |
| | 60-69 | **1728** | **19%** | 284 (19%) | 300 (20%) | 282 (19%) | 312 (21%) | 276 (18%) | 274 (18%) |
| | 70-79 | **518** | **6%** | 66 (4%) | 90 (6%) | 103 (7%) | 95 (6%) | 77 (5%) | 87 (6%) |
| | 80+ | **40** | **0%** | 3 (0%) | 6 (1%) | 10 (1%) | 7 (0%) | 10 (1%) | 4 (0%) |
| Sex | | | | | | | | | |
| | Female | **4250** | **47%** | 719 (48%) | 699 (47%) | 669 (45%) | 725 (48%) | 728 (49%) | 710 (47%) |
| | Male | **4749** | **53%** | 781 (52%) | 800 (53%) | 831 (55%) | 775 (52%) | 772 (51 %) | 790 (53%) |

**Supplemental Table 2. Assay Characteristics overall and over time; based on the BLCM Posterior Means (95% Creditable Interval), Informative**

| | Overall | | | | April/May | | June/July | | August/September | |
|---|---|---|---|---|---|---|---|---|---|---|
| | Sensitivity | PPV | Specificity | NPV | Sensitivity | Specificity | Sensitivity | Specificity | Sensitivity | Specificity |
| Spike | 93.5% (88.7, 97.3) | 27.7% (23.2, 33.7) | 98.1% (97.9, 98.4) | 99.8% (99.6, 100.0) | 95.0% (90.1, 98.2) | 98.0% (97.4, 98.4) | 93.6% (88.0, 97.6) | 98.8% (98.4, 99.2) | 93.6% (88.0, 97.6) | 97.6% (97.0, 98.1) |
| RBD | 89.1% (84.1, 93.5) | 59.8% (50.0, 71.4) | 99.5% (99.3, 99.7) | 99.8% (99.6, 99.9) | 89.3% (83.7, 93.8) | 99.5% (99.3, 99.8) | 89.2% (98.4, 99.2) | 99.6% (83.6, 93.9) | 88.7% (83.0, 93.4) | 99.3% (99.0, 99.6) |
| NP | 78.8% (74.1, 83.2) | 21.0% (17.2, 25.3) | 98.2% (97.9, 98.4) | 99.6% (99.3, 99.7) | 79.9% (74.9, 84.5) | 99.5% (99.2, 99.7) | 80.5% (75.6, 84.9) | 97.3% (96.7, 97.9) | 78.5% (73.5, 83.3) | 97.6% (97.1, 98.1) |
| Abbott-NP | 58.5% (46.3, 70.6) | 87.5% (81.3, 93.8) | 99.8% (99.7, 99.9) | 99.4% (99.1, 99.6) | 77.3% (58.7, 92.5) | 99.7% (99.5, 99.9) | 60.2% (41.2, 78.5) | 99.8% (99.5, 99.9) | 64.4% (45.6, 83.0) | 99.9% (99.8, 100.0) |

**Supplemental Table 3. Assay Characteristics overall and over time; based on the BLCM Posterior Means (95% Creditable Interval), Weakly Informative**

| | Overall | | | | April/May | | June/July | | August/September | |
|---|---|---|---|---|---|---|---|---|---|---|
| | Sensitivity | PPV | Specificity | NPV | Sensitivity | Specificity | Sensitivity | Specificity | Sensitivity | Specificity |
| Spike | 92.9% (83.9, 99.4) | 27.1% (24.6, 29.8) | 98.1% (97.8, 98.4) | 99.9% (99.9, 100.0) | 96.7% (87.2, 100.0) | 97.9% (97.3, 98.4) | 92.0% (76.9, 99.8) | 98.7% (98.3, 99.1) | 92.2% (77.2, 99.8) | 97.5% (96.9, 98.0) |
| RBD | 90.6% (80.5, 98.1) | 57.8% (51.9, 62.5) | 99.5% (99.3, 99.6) | 99.9% (99.9, 100.0) | 91.3% (77.0, 99.0) | 99.4% (99.1, 99.7) | 93.2% (77.9, 99.9) | 99.4% (99.1, 99.7) | 89.7% (73.5, 99.5) | 99.2% (98.8, 99.5) |
| NP | 75.8% (64.6, 85.9) | 23.0% (20.6, 25.7) | 98.1% (97.8, 98.4) | 99.8% (99.8, 99.9) | 84.8% (66.6, 97.6) | 99.4% (99.1, 99.6) | 89.4% (74.1, 99.3) | 97.2% (96.6, 97.8) | 70.5% (52.1, 88.0) | 97.5% (96.9, 98.1) |
| Abbott-NP | 58.7% (46.2, 71.2) | 67.2% (64.8, 70.4) | 99.8% (99.7, 99.9) | 99.7% (99.6, 99.8) | 79.4% (59.8, 94.5) | 99.6% (99.4, 99.8) | 62.9% (42.4, 82.1) | 99.6% (99.4, 99.8) | 63.9% (44.1, 83.9) | 99.8% (99.6, 99.9) |
